# Proteomic Profiling of MIS-C Patients Reveals Heterogeneity Relating to Interferon Gamma Dysregulation and Vascular Endothelial Dysfunction

**DOI:** 10.1101/2021.04.13.21255439

**Authors:** Caroline Diorio, Rawan Shraim, Laura A. Vella, Josephine R. Giles, Amy E. Baxter, Derek A. Oldridge, Scott W. Canna, Sarah E. Henrickson, Kevin O. McNerney, Frances Balamuth, Chakkapong Burudpakdee, Jessica Lee, Tomas Leng, Alvin Farrell, Michele P. Lambert, Kathleen E. Sullivan, E. John Wherry, David T. Teachey, Hamid Bassiri, Edward M. Behrens

## Abstract

Multi-system Inflammatory Syndrome in Children (MIS-C) is a major complication of the Severe Acute Respiratory Syndrome Coronavirus 2 (SARS-CoV-2) pandemic in pediatric patients. Weeks after an often mild or asymptomatic initial infection with SARS-CoV-2 children may present with a severe shock-like picture and marked inflammation. Children with MIS-C present with varying degrees of cardiovascular and hyperinflammatory symptoms. We performed a comprehensive analysis of the plasma proteome of more than 1400 proteins in children with SARS-CoV-2. We hypothesized that the proteome would reflect heterogeneity in hyperinflammation and vascular injury, and further identify pathogenic mediators of disease. Protein signatures demonstrated overlap between MIS-C, and the inflammatory syndromes macrophage activation syndrome (MAS) and thrombotic microangiopathy (TMA). We demonstrate that PLA2G2A is a key marker of MIS-C that associates with TMA. We found that IFNγ responses are dysregulated in MIS-C patients, and that IFNγ levels delineate clinical heterogeneity.

---

Multisystem inflammatory syndrome in children (MIS-C) emerged as the major pediatric complication of the Severe Acute Respiratory Syndrome Corona Virus 2 (SARS-CoV-2) pandemic of 2020.^1–3^ MIS-C is characterized by fever and inflammation, and has some clinical features overlapping with Kawasaki disease (KD).^3–7^ Due to the similarities with KD, initial treatment regimens for MIS-C included both intravenous immunoglobulin (IVIG) and corticosteroids.^8–10^ Data from randomized controlled trials comparing these two treatments is not yet available, however, best evidence to date suggests that IVIG alone provides suboptimal treatment for MIS-C.^8^

Although the phenotypes of KD and MIS-C are overlapping, they have important clinical differences and immune profiling has delineated separate but overlapping signatures between the conditions.^5, 11–13^ Shock is a marked feature in patients with MIS-C, with the majority of patients requiring pediatric intensive care unit (PICU) admission.^4^ The severity of MIS-C has invoked cytokine storm as a potential driver of symptoms.^11^ SARS-CoV-2 infection leading to severe Corona Virus Disease (COVID-19) can also lead to cytokine storm.^14^ Multiple cytokines including interferon gamma (IFNγ), interleukin-6 (IL-6), IL-8, IL-1, IL-17 and IL-1β have been implicated as potentially causative in patients with MIS-C.^11, 15^ The putative role of cytokine dysregulation in the pathophysiology of MIS-C has led to the use of cytokine blockade with agents such as tocilizumab or anakinra in refractory patients, however, the optimal targeted agent has not yet been determined.^9^ Improved understanding of the cytokine dysregulation in these patients is critical to identifying rational targeted therapies.

In addition to shock and inflammation, MIS-C and SARS-CoV-2 infection are associated with an increased risk of thrombosis in both adult and pediatric patients (Whitworth et al. *Blood*, in press).^16^ We have previously demonstrated that infection with SARS-CoV-2 and MIS-C are associated with thrombotic microangiopathy (TMA), and the associated biomarker soluble C5B9 (SC5B9).^17^ TMA is a group of diseases characterized by microangiopathic hemolytic anemia, thrombocytopenia and organ dysfunction related to microthrombi.^18^ TMA can occur as a primary process or secondary to infection, inflammatory insult or in hematopoietic stem cell transplant (HSCT).^18, 19^ The mechanism of TMA in the context of MIS-C is not known, and its association with cytokine dysregulation remains unclear.

We interrogated >1400 proteins in the plasma proteome and integrated this information with clinical and high-dimensional flow cytometry data. We hypothesized that distinct dysregulations in proteins and associated pathways that relate to cytokine storm and vascular injury contribute to the underlying pathophysiology of MIS-C. Our goals were to understand the pathophysiology of MIS-C via a thorough examination of the plasma proteome, and to identify candidate biomarkers and predictors of disease severity.

## RESULTS

### Patients Included in the Study

Between April 2020 and October 2020, we enrolled 63 hospitalized patients with MIS-C (N=22), Severe COVID-19 (“Severe”, N=15) or asymptomatic or mild SARS-CoV-2 infection (“Minimal”, N=26). In addition, we included remnant plasma samples from otherwise healthy patients (“Healthy”, N=25). Details of the clinical presentation of MIS-C, Severe, and Minimal patients are presented in Supplemental Tables S1 and S2. Similar to previously reported cohorts, a high proportion (N = 17, 77%) of patients with MIS-C were admitted to the PICU.^2, 4^ We utilized the Olink Explore 1536/384 protein biomarker platform to interrogate the plasma proteome of all 88 patients. A subset of patients included in this study have been reported previously.^17, 20–23^ Flow cytometry data was available for 20 patients included in this study (MIS-C N=8, Severe N=6, Minimal N=6), reported previously by Vella et al.^20^

We first sought to validate the accuracy of the O-link platform. Patients with MIS-C in our sample had marked elevations of brain-type natriuretic peptide (BNP) and troponin measured clinically (Supplemental Table S2). The Olink panel measures N-terminal prohormone brain natriuretic peptide (NTproBNP), a protein pre-cursor to BNP associated with cardiac damage.^24^ NTproBNP was significantly higher in MIS-C patients (Supplemental Figure S1A), consistent with other reports.^15^ As expected, Olink NTproBNP correlated highly with clinical BNP (R=0.78, p =2.7e-08; Supplemental Figure S1B) .

To further validate the data, we compared cytokines measured in our laboratory with the same cytokines measured by Olink and demonstrated strong correlations (Supplemental Figures S1C). We and others have previously found elevations in IL-10 distinguish patients with MIS-C from those with acute SARS-CoV-2 infection. We confirmed this observation with data from Olink (Supplemental Figure S1D).^13, 21^

### Overall Architecture of Pediatric SARS-CoV-2 and MIS-C Plasma Proteome

Following this validation, we next took an unbiased approach to investigate the proteomic data. The overall architecture of the data is presented in Figure 1. Patients with MIS-C cluster differently from patients with acute SARS-CoV-2 or healthy patients when visualized with t-istributed stochastic neighbor embedding (tSNE; Figure 1A). Vella et al. previously demonstrated that MIS-C is associated with CD8+ T-cell activation and Tbet+ plasmablasts.^20^ In the subset of patients on whom flow cytometry data was available, we overlaid flow cytometry markers on tSNE clustering. Interestingly, there was heterogeneity in percent of activated CD8+ T-cells and percent of Tbet+ plasmablasts within the MIS-C clusters (Figure 1A).^20^ Furthermore, there appears to be an inverse relationship between regions with higher percentage of activated CD8+ cells and a higher percentage Tbet+ plasmablasts within these MIS-C subclusters.

**Figure 1.**
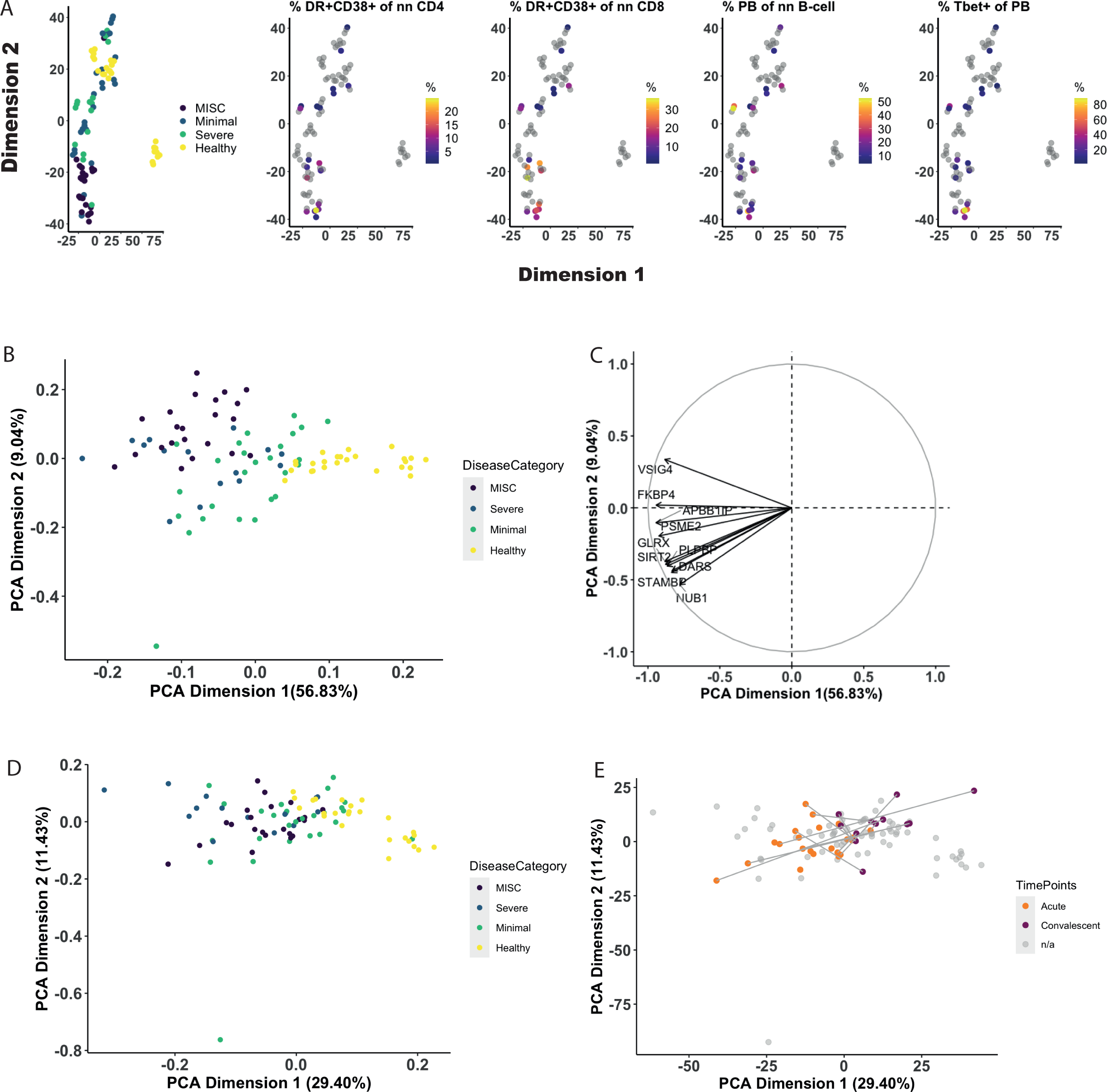
Overarching architecture of plasma proteome in patients with Multisystem Inflammatory Syndrome in Children (MIS-C; N=22), minimal Severe Acute Respiratory Syndrome Corona Virus 2 (SARS-CoV-2; N=26) infection, and Severe Coronavirus Disease (COVID-19; N=15) compared to healthy controls (N=25). t-distributed Stochastic Neighbor Embedding (tSNE) plots are used to visualize clustering between the four groups of patients (A) with overlay from flow cytometry-based scores of percent of non-naïve CD4+ T-cells that are activated (HLA-DR+CD38+), percent non-naïve CD8+ T-cells that are activated, percent of non-naive B-cells that are plasmablasts, and percent of plasmablasts that are T-bet+. Gray dots indicate that data was not available. Clustering was further examined using principal component analysis (PCA) of DEPs between all 4 patient groups, log2 fold-change threshold of 2 and FDR threshold of 0.01 (B). The top 20 proteins contributing to PCA Dimensions 1 and 2 are presented in Panel C. To understand the change in the proteome over time in MIS-C patients we first created a PCA of all proteins for all disease categories (D). MIS-C patients with matched acute and convalescent samples (N=12) were then transformed onto the space. Acute samples are shown in orange and convalescent samples in purple. Lines connect matched pairs.

In order to look at loading plots of the most prominent drivers of difference between groups, we used principal component analysis (PCA) to visualize clustering between patients. To achieve better separation between groups, we limited the analysis to proteins that were differentially expressed between any of the four groups (N=231). Lists of differentially expressed proteins (DEP) between all groups are available in Supplemental Material M1. The corresponding Scree plot and top 20 contributions to each dimension of the PCA are available in Supplemental Figures S2A-C. Multiple IFNγ responsive proteins contribute to PCA Dimension 2, including IL-27, CXCL9, IL18BP and CCL23. Healthy patients clustered separately from those with Severe, Minimal or MIS-C (Figure 1B), while patients with Severe and MIS-C overlapped. The top ten contributing proteins that account for the variance associated with PCA Dimensions 1 and 2 are presented in Figure 1C.

Twenty patients with MIS-C received treatment with both steroids and IVIG. One patient was treated with IVIG alone, and one patient did not receive treatment. Convalescent samples were available on 12 patients (54%). To understand how the overall proteome of patients with MIS-C changes over time, we first created a PCA mapping all proteins and all patients (Figure 1D). We then PCA transformed data for convalescent samples on that space (Figure 1E). Convalescent samples shifted towards the healthy controls, implying that following treatment, the proteome in MIS-C patients returns towards a baseline state. Pre-and post-treatment samples were available on 5 patients. To investigate if the timing of initial collection relative to treatment altered the proteome, we assessed change over time in one of the most differentially expressed proteins, phospholipase A2 (PLA2G2A; Supplemental Figure S1E). PLA2G2A pre-and post-treatment samples did not significantly differ in acute timepoints suggesting that sampling of plasma shortly after treatment does not immediately affect results.

### Uncovering Pathways of Activation in Patients with MIS-C and SARS-CoV-2 Infection

We used unbiased exploration of DEPs to understand pathways of activation between MIS-C, Minimal and Severe patients compared to healthy patients. Volcano plots are presented in Figure 2. Notably, PLA2G2A is highly differentially expressed between all three groups compared to healthy patients, with the most marked difference occurring in MIS-C patients.

**Figure 2.**
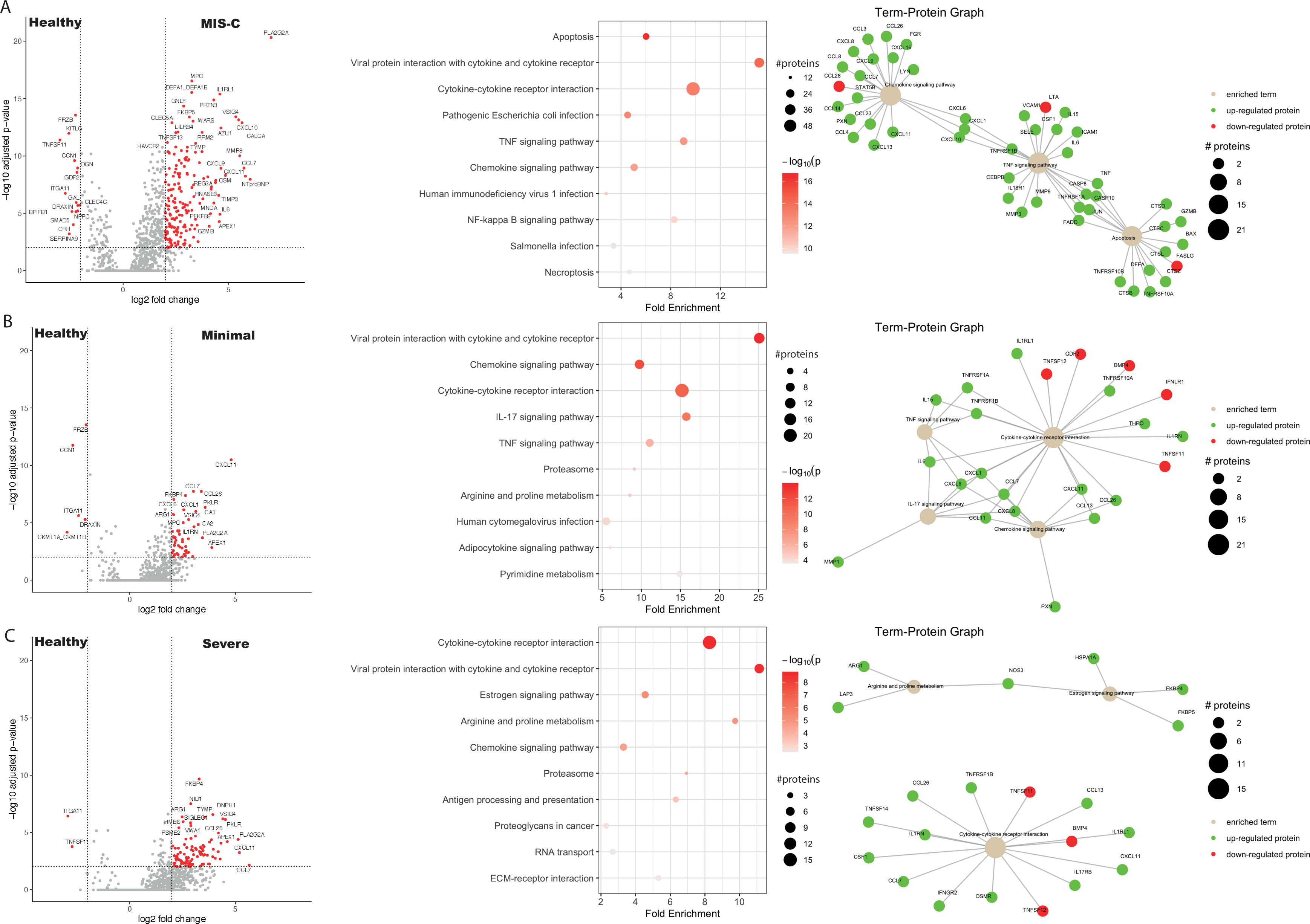
Differentially expressed proteins, ranked pathway analysis and network analysis for each disease state compared to healthy controls. Differentially expressed proteins, log2 fold change threshold of 2 and FDR threshold of 0.01, between patients with MIS-C (N=22) and healthy controls (N=25) are shown in (A), along with ranked pathway analysis. Size of dots represents number of enriched genes and intensity of colors represents p-value associated with enrichment analysis. Network analyses are shown where up-regulated proteins are colored in green and down-regulated proteins are colored in red. Node size of the pathway term represents number of input proteins implicated in the pathway. The same series of graphs is shown for patients with minimal Severe Acute Respiratory Syndrome Corona Virus 2 (SARS-CoV-2; N=26) infection compared healthy controls (B), and Severe Coronavirus Disease (COVID-19; N=15) patients compared to healthy controls (C).

In order to identify pathways of relevance to the pathophysiology of MIS-C and pediatric COVID-19 we performed pathway analysis accounting for protein-protein interactions using pathfindR.^25^ PathfindR uses an active subnetwork enrichment analysis approach to identify lists of enriched pathways. Ranked pathways and enrichments are presented in Figure 2A-C. Lists of all ranked pathways are available in Supplemental Material S2. A protein to pathway analysis on the pathways of interest was performed to identify interactions between various proteins in the datasets and pathways.

Cytokine-cytokine and chemokine-cytokine receptor pathways were dysregulated in all classifications of pediatric SARS-CoV-2 infection. Notably, similar pathways were perturbed in MIS-C, Minimal and Severe patients compared to healthy controls. Overlaps of DEPs between disease states are shown in Supplemental Figure S3A. The majority of DEPs in Severe COVID-19 are also differentially expressed in MIS-C. We directly assessed the DEPs between these groups and surprisingly, there were very few differences (Supplemental Figure S3B). As this comparison involved a smaller proportion of patients, we used a nominal p-value with a threshold of 0.05.

### MIS-C Is Characterized by a Disproportionate Response to Interferon Gamma

IFNγ has been associated with MIS-C in previous reports.^12, 15^ We previously described that IFNγ levels were high in patients with Severe and MIS-C, that levels of IFNγ could not distinguish between these two groups and that IL-10 can distinguish MIS-C from Severe (Supplemental Figure S3B).^21^ In order to better understand these two cytokines, we investigated correlations between IFNγ and IL-10 and their canonical response proteins as well as proteins associated with cellular populations responsible for their production (Figure 3). We examined CXCL9 as the key protein associated with IFNγ response and found that all patients with SARS-CoV-2 infection showed a positive correlation between CXCL9 and IFNγ. Strikingly, MIS-C patients had a disproportionately high CXCL9 response to IFNγ compared to the other groups (Figure 3A; p<0.001 by logistical regression modelling). To probe the source of IFNγ production, we correlated IFNγ levels with soluble markers of activated T-cells (IL2RA), NK-cells (NCR1) and macrophages (CD163). IFNγ levels in MIS-C patients associated significantly with IL2RA and NCR1 but not CD163, suggesting that IFNγ levels correlate with T-cells and NK-cells but not macrophages (Figure 3A).

**Figure 3.**
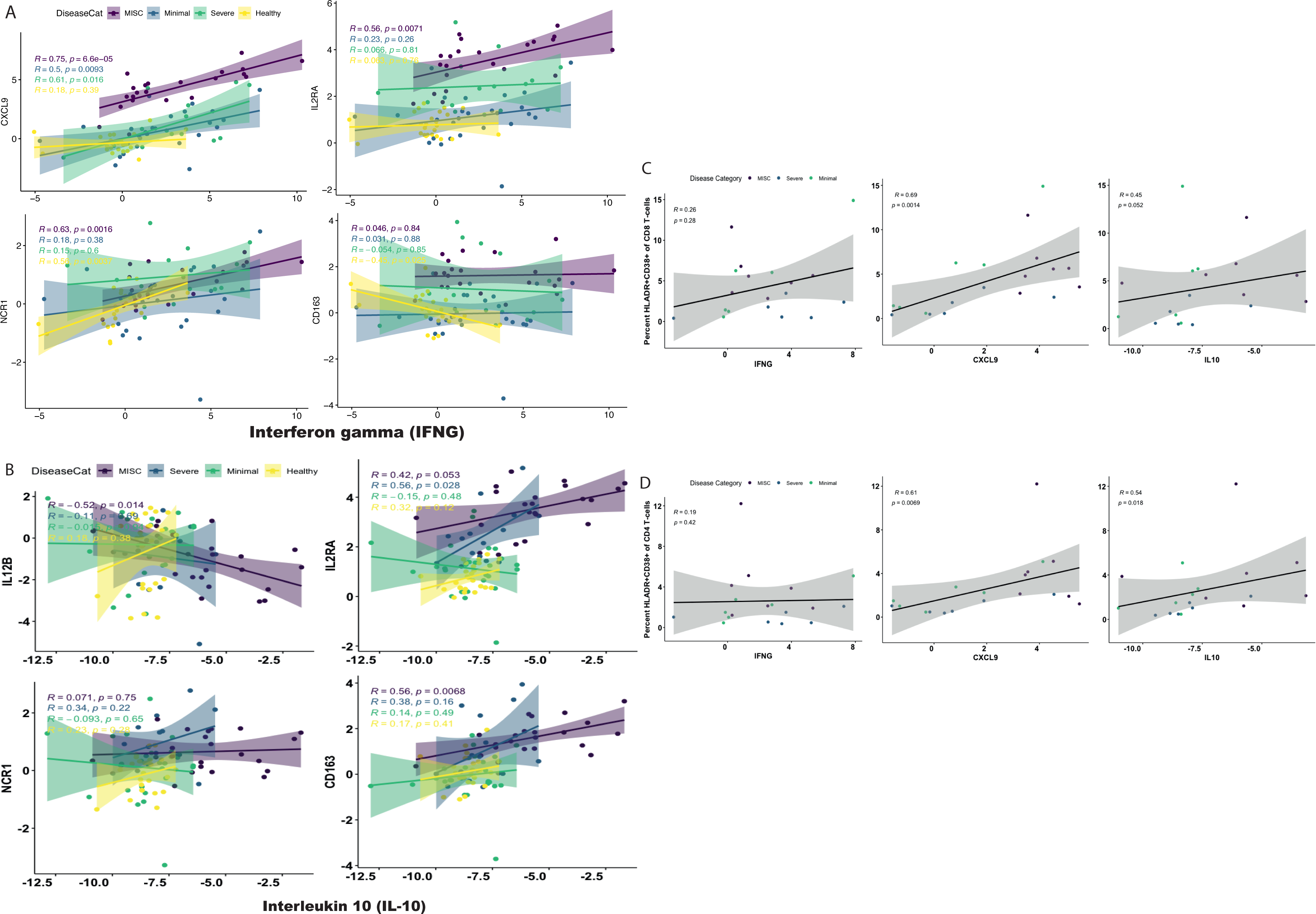
Interferon gamma (IFNγ) and interleukin-10 (IL-10) responses and associated cell types. Spearman correlations between IFNγ signaling and its canonical response protein (CXCL9), as well as proteins associated with activated T-cells (IL2RA), NK cells (NCR1) and macrophages (CD163) for patients with (MIS-C; N=22), Minimal Severe Acute Respiratory Syndrome Corona Virus 2 infection (SARS-CoV-2; N=26) infection, and Severe Corona Virus Disease (COVID-19; N=15) compared to healthy controls (N=25) are shown in panel (A). Correlations between IL-10 and its canonical receptor IL12B, as well as IL2RA, NCR1 and CD163 for each disease category are shown in panel (B). Panel (C) shows correlations between HLADR+CD38+ CD8+ T-cells (N=19) and IFNγ, CXCL9, and IL-10. Dots are colored by each patient’s disease category. Panel (D) demonstrates correlations between HLADR+CD38+ CD4+ T-cells (N=19) and IFNγ, CXCL9, and IL-10. Dots are colored by disease category.

Levels of IL-10 have been shown previously, and in this cohort, to be significantly higher in patients with MIS-C than in those with Severe COVID-19 (Supplemental Figure S1D).^13, 21^ Similar to the analysis above, we examined the expression of IL12B, which should be inhibited by IL-10.^26^ IL12B was inversely correlated with IL-10 in patients with MIS-C, but not in Minimal, Severe or Healthy patients (Figure 3B). This finding implies that the elevated IL-10 in MIS-C patients is bioactive. IL-10 production was significantly associated with markers of activated T-cells and macrophages, but not with NK-cells (Figure 3B).

We subsequently examined the relationship between IFNγ, CXCL9 and IL-10 and activated CD8+ and CD4+ T-cells from flow cytometry data in order to probe these correlations further (Figure 3C&D). We found that CXCL9 was significantly and strongly correlated with both cell types but IFNγ was not, suggesting T-cells are being activated by IFNγ rather than producing IFNγ. IL-10 was significantly associated with activated CD4+ but not activated CD8+ T-cells.

### MIS-C Patients have an MAS-like phenotype characterized by IFN**γ** and CXCL9 signaling

The outsized response to IFNγ with excessive CXCL9 expression in patients with MIS-C evoked the cytokine phenotype of macrophage activation syndrome (MAS).^27, 28^ MAS is a hyperinflammatory condition that occurs in the setting of rheumatologic diseases, infections or as a primary disorder, characterized by excessive activation of lymphocytes and macrophages in the setting of marked IFNγ production.^29, 30^ We therefore investigated whether IFNγ, CXCL9 and other markers associated with MAS could distinguish different SARS-CoV-2 outcomes (Figure 4A).^28, 30, 31^ Unsupervised hierarchical clustering revealed two distinct groups of MIS-C patients that had elevations in most of the MAS markers. These two clusters primarily differed by their IFNγ expression with an IFNγ-high versus IFNγ-low group evident.

**Figure 4.**
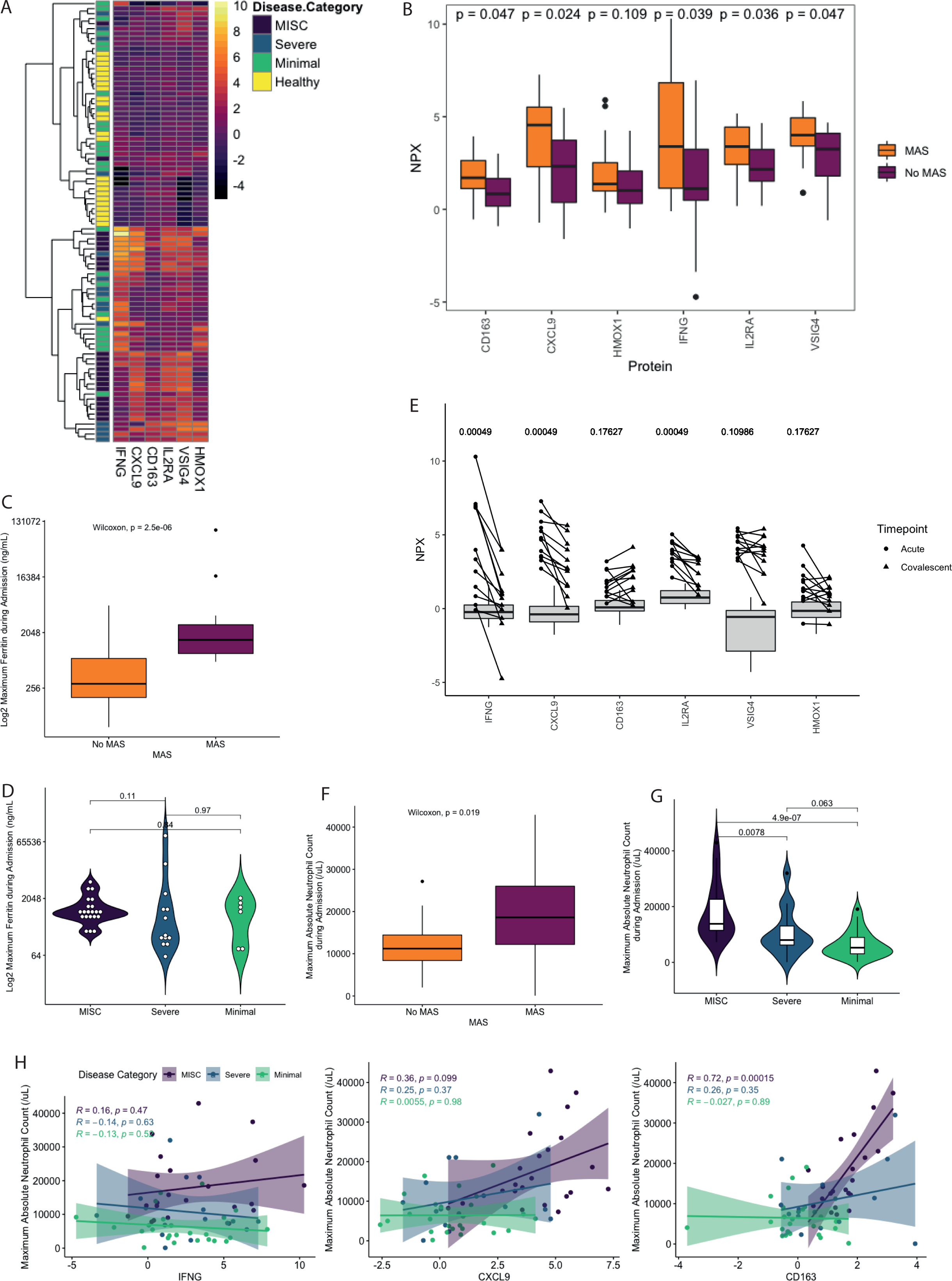
Association with macrophage activation syndrome (MAS) and patients with Multisystem Inflammatory Syndrome in Children (MIS-C) and Severe Acute Respiratory Syndrome Corona Virus 2 (SARS-CoV-2) infection. (A) Heatmap of MAS-associated proteins IFNγ, CXCL9, CD163, IL2RA, VSIG4 and HMOX1 with unsupervised hierarchical clustering applied to all patients (N=88). (B) Boxplot demonstrating levels of MAS-associated proteins in patients who met (N=17) and did not meet modified criteria for MAS (N=23). P-values computed with Wilcoxon test. (C) Median log2 transformed maximum ferritin during admission in patients who did (N=17) and did not (N=23) meet criteria for MAS. P-value computed with Wilcoxon test. (D) log2 transformed maximum ferritin during admission for patients with MIS-C (N=21), Severe COVID-19 (N=13) and minimal disease (N=6). P-values computed with pairwise comparisons using Wilcoxon rank sum test following Kruskal-Wallis testing. (E) Acute and convalescent levels of each MAS-related protein for MIS-C patients on whom both acute and convalescent samples were available (N=12). Circles represent acute samples with triangles representing convalescent samples. Lines connect matched pairs. Gray boxes and whiskers show median and interquartile range of healthy controls (N=25). P-values for difference between acute and convalescent samples were computed using a paired samples Wilcoxon test. (F) Boxplot of maximum absolute neutrophil count during admission for patients who met (N=17) and did not meet (N=23) criteria for MAS. P-value computed with Wilcoxon test. (G) Maximum absolute neutrophil count during admission for patients with MIS-C (N=22), Severe disease (N=15) and Minimal disease (N=26). P-values computed with pairwise comparisons using Wilcoxon rank sum test following Kruskal-Wallis testing. (H) Maximum absolute neutrophil count during admission correlated with IFNγ, CXCL9 and CD163. R computed using Spearman correlation.

To further interrogate the hypothesis that MAS was associated with MIS-C, we applied a modified version of the Ravelli MAS criteria to all patients on whom ferritin had been measured (N=40; MIS-C N=21, Minimal N=6, Severe N=13).^32^ If parameters were missing, they were imputed as negative to bias our score towards the null hypothesis. We compared patients who met criteria for MAS to those who did not (Supplemental Table S3). CD163, CXCL9, IFNγ, IL2RA and VSIG4 were significantly elevated in patients who met MAS criteria (Figure 4B).

We found that maximum ferritin during admission was significantly higher in patients that met criteria for MAS than those that did not (P<0.0001) but did not differ significantly between the disease groups as a whole (Figure 4C&D). In MIS-C patients on whom both an acute and a convalescent sample was available (N=12), IFNγ, CXCL9 and IL2RA all significantly decreased over time (Figure 4E). For patients with MIS-C, time of sample draw relative to treatment with IVIG or steroids against IFNγ level is presented in Supplemental Figure S1F and shows decay over time.

MAS is typically associated with cytopenias, including neutropenia. In contrast, MIS-C is characterized by marked neutrophilia,^4^ and in patients who met criteria for MAS high absolute neutrophil counts (ANC) were apparent (Figure 4F). ANC was significantly higher in patients with MIS-C than in those with Severe or Minimal (Figure 4G). In MIS-C patients, neutrophilia did not correlate with IFNγ or CXCL9 but correlated strongly with CD163, implying neutrophilia in these patients is associated with macrophage activation (R=0.72, p<0.0001; Figure 4H), but not directly related to IFNγ or CXCL9. Macrophage hyperresponsiveness to IFNγ expression has been linked to TRIM8 dysregulation in MAS.^33^ We noted dysregulation of another TRIM protein, the IFNγ signaling suppressive protein TRIM21.^34, 35^ Consistent with a derepression of IFNγ signaling, MIS-C patients expressed less TRIM21 compared to healthy controls (Supplemental Figure S3C).

### PLA2G2A is a candidate biomarker for MIS-C and is associated with a thrombotic microangiopathy phenotype

We and others have previously demonstrated that infection with SARS-CoV-2 is associated with clinical TMA and elevations in the TMA biomarker SC5B9.^17, 36^ As SC5B9 was not included in the Olink analysis, we correlated SC5B9 levels measured in our laboratory (N=75) to Olink proteins related to vascular or platelet dysfunction to identify surrogate biomarkers. Hierarchical clustering identified that SC5B9 correlated most highly with PLA2G2A, PDGFC, SELE, CALCA, NOS3, VWA1 and TYMP (Figure 5A).

**Figure 5.**
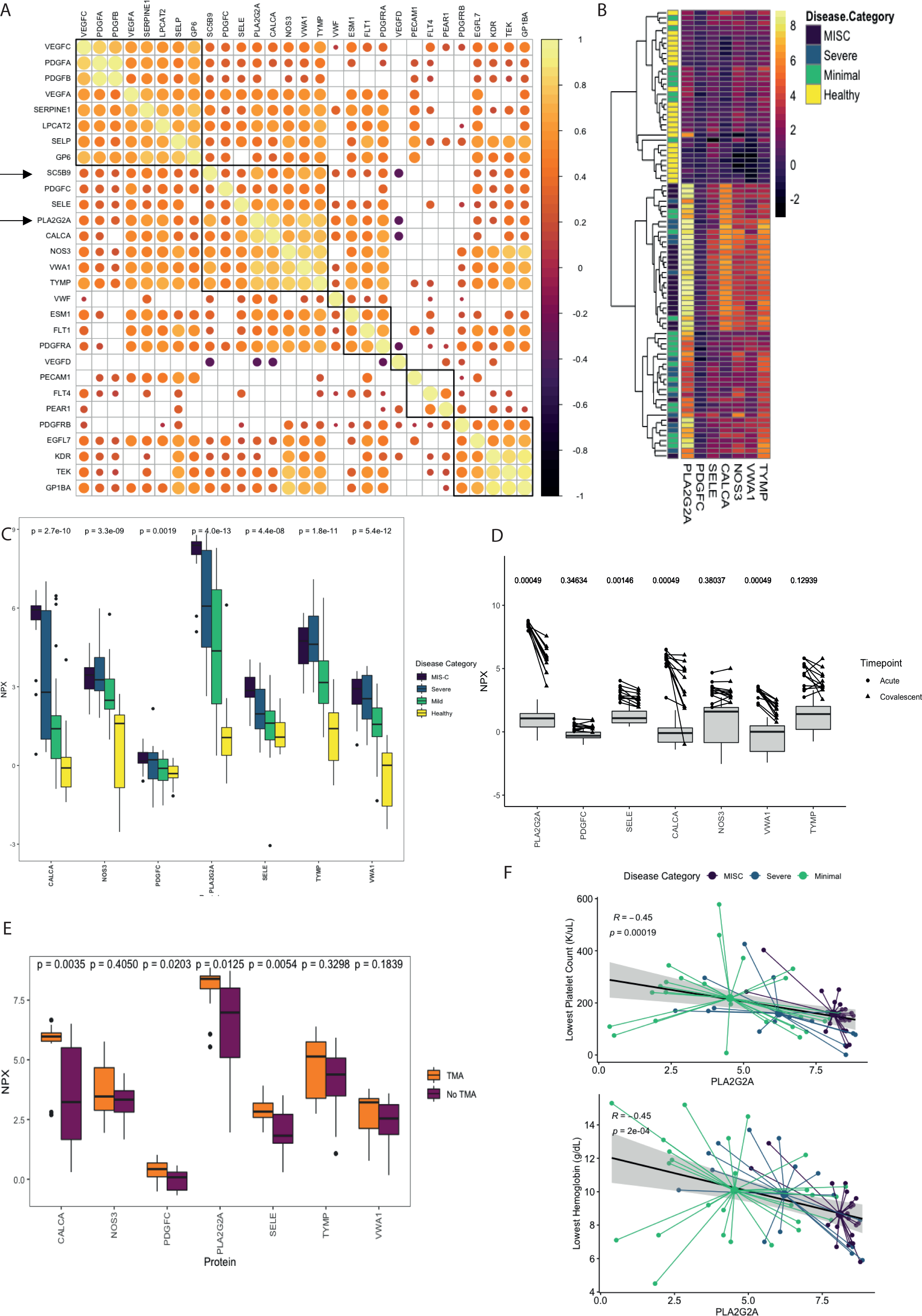
Evidence of thrombotic microangiopathy and vascular endothelial dysfunction in patients with Multisystem Inflammatory Syndrome in Children (MIS-C) and Severe Acute Respiratory Syndrome Corona Virus 2 (SARS-CoV-2) infection. (A) Correlation matrix of vascular and platelet related proteins and soluble C5B9 (SC5B9) in all patients on whom an SC5B9 was measured (N=75). Hierarchical clustering was applied to identify surrogate markers associated with SC5B9. (B) Heatmap of candidate surrogate markers associated with SC5B9 including PLA2G2A, PDGFC, SELE, CALCA, NOS3, VWA1 and TYMP. Unsupervised hierarchical clustering was applied to all patients (N=88) with boxes colored by disease category. (C) Boxplots of each SC5B9 related protein across disease states. P-values computed by Kruskal-Wallis testing. (D) Acute and convalescent levels of each SC5B9-related protein for MIS-C patients on whom both acute and convalescent samples were available (N=12). Circles represent acute samples with triangles representing convalescent samples. Lines connect matched pairs. Gray boxes and whiskers show median and interquartile range of healthy controls (N=25). P-values for difference between acute and convalescent samples were computed using a paired samples Wilcoxon test. (E) Boxplot demonstrating levels of SC5B9-associated proteins in patients who met (N=13) and did not meet criteria for thrombotic microangiopathy (TMA; N=21). P-values computed with Wilcoxon test. (F) Correlations between lowest platelet count and lowest hemoglobin during admission and PLA2G2A. Small circles represent individual patients with large central circles representing the mean for each disease category. Dots are colored by disease category as MIS-C (N=22), Severe (N=15) or Minimal (N=26). R computed with Spearman correlation.

We then evaluated whether proteins highly correlated with SC5B9 were able to identify disease category (Figure 5B). Patients with MIS-C clustered due to very high PLA2G2A expression, and moderate CALCA and TYMP expression. PLA2G2A and CALCA were both significantly higher between MIS-C and Severe patients (Figure 5C). PLA2G2A is significantly higher in SARS-CoV-2 infected patients than in healthy patients with the most marked difference in patients with MIS-C (Figure 5C). In MIS-C patients, PLA2G2A, SELE, CALCA and VWA1 all improved between the acute and convalescent phases (Figure 5D). Notably, levels of PLA2G2A do not return completely to normal during convalescence.

For patients who had blood smears available for review (N=34; MIS-C N=15, Minimal N=11, Severe N=8) we applied simple criteria for TMA, as previously published (Supplemental Table S4).^17^ Meeting criteria for TMA was associated with a significantly higher PLA2G2A and CALCA (Figure 5E). Patients who met criteria for TMA were more likely to require inotropes during their admission than those who did not meet criteria for TMA (92% vs 23% respectively, N=34, p-value <0.001). Given our hypothesis that PLA2G2A is a marker of TMA and microangiopathic hemolytic anemia, we examined whether levels of PLA2G2A correlated to lowest platelet count and lowest hemoglobin during admission. PLA2G2A levels inversely correlated with platelets and hemoglobin with groupings by disease category evident (Figure 5F).

### Clinical heterogeneity among MIS-C Patients

Next, we investigated the intersection between the MAS and TMA phenotypes and MIS-C. Among MIS-C patients, TMA and MAS occurred independently (Fisher’s exact, p = n.s.; Figure 6A). Furthermore, when TMA and MAS phenotypes were overlayed on the same tSNE mapping used in Figure 1, patient clustering by disease state was evident (Supplemental Figure S4A). To further explore the heterogeneity among MIS-C patients, we compared patients from the IFNγ-high (N=7) and IFNγ-low (N=13) clusters identified in Figure 4A. We looked at DEPs between these two clusters and performed unbiased pathway and clustering analysis (Figure 6B-D) revealing differences in cytokine signaling including IL-17 pathways. IL-17 is a neutrophil chemotactic factor and may relate to the neutrophilia present in these patients.^37^ In Supplemental Figure S4B IFNγ levels are overlayed on the same tSNE map as displayed in Figure 1, demonstrating heterogenous clustering within MIS-C.

**Figure 6.**
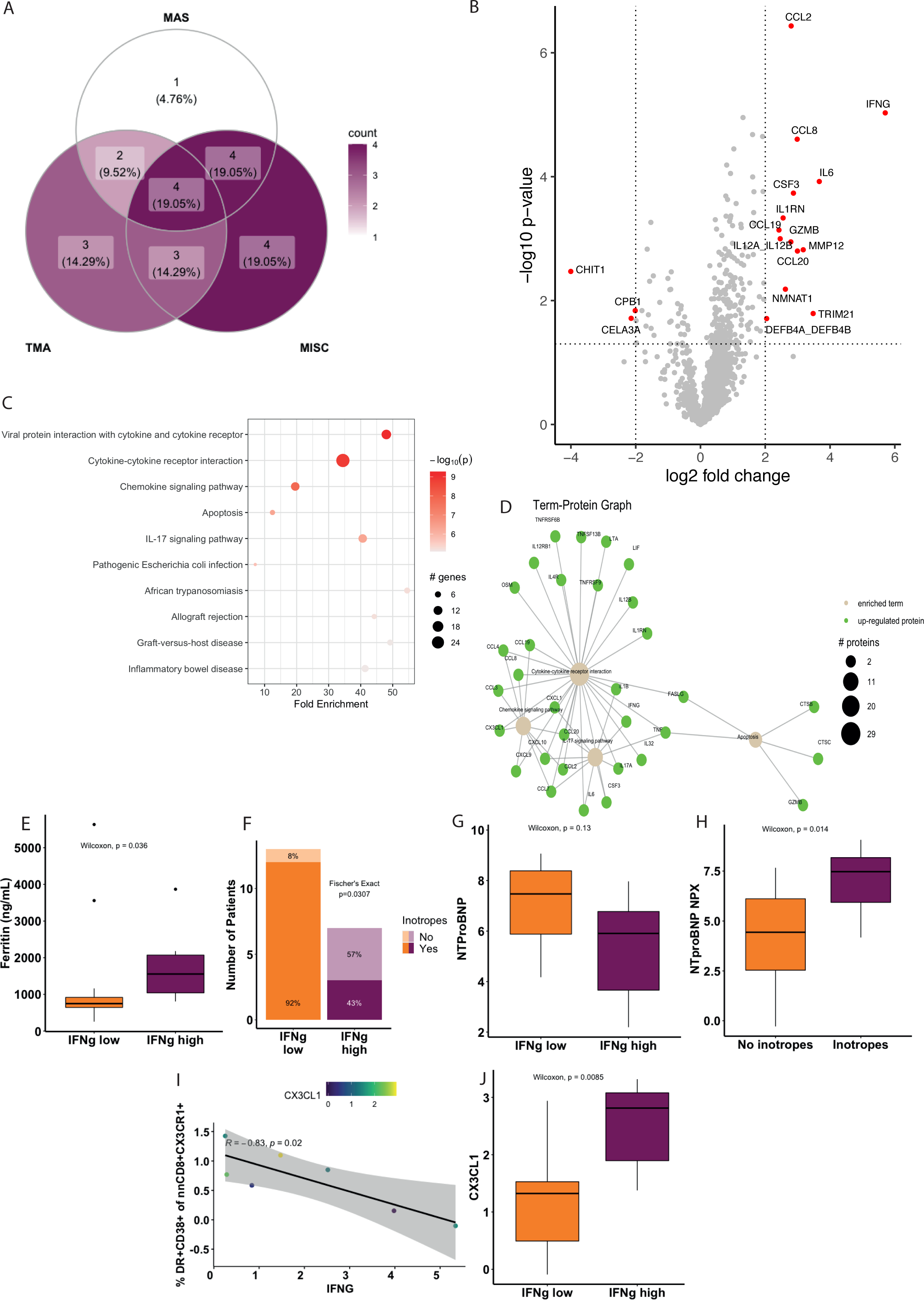
Clinical heterogeneity among Multisystem Inflammatory Syndrome in Children (MIS-C) patients defined by their interferon gamma (IFNγ) signature. (A) Overlaps between patients who meet criteria for macrophage activation syndrome (MAS), thrombotic microangiopathy (TMA) and MIS-C. (B) Differentially expressed proteins between patients with IFNγ-low (N=13) and IFNγ-high expression (N=7). Log2fold change threshold of 2 and a nominal p-value cutoff of 0.05 were used. (C) Unsupervised pathway ranking and (D) network analysis for differentially expressed proteins between IFNγ-low and IFNγ-high patients. (E) Maximum ferritin level during admission for patients in the IFNγ-low and IFNγ-high. P-value computed using Wilcoxon test. (F) Number of patients in IFNγ-low and IFNγ-high groups who did and did not require inotropic support. P-value computed with Fisher’s exact test. NTproBNP expression between IFNγ-low and IFNγ-high groups (G) and in MIS-C patients who did (N=7) and did not require inotropes (H; N=15). (I) Correlation between IFNγ level and percent DR+CD38+ non-naïve CD8+ CX3CR1+ T-cells in MIS-C patients (N=7). Dots colored by CX3CL1 expression. R value computed using Pearson’s correlation coefficient after normality was demonstrated. (J) CX3CL1 levels between IFNγ-low and IFNγ-high groups. P-values computed with Wilcoxon test.

We examined occurrence of TMA and MAS among these IFNγ-high and -low MIS-C clusters. There was no increase in TMA among IFNγ-high MIS-C patients (Fisher’s exact, p = n.s.; Supplemental Table S5). There was a trend towards a significantly increased enrichment in MAS among IFNγ-high MIS-C patients compared to IFNγ-low patients (83% vs. 38%, Fisher’s exact p=0.14; Supplemental Table S5). In keeping with this trend, IFNγ-high MIS-C patients had significantly higher ferritin (p=0.036; Figure 6E). IFNγ status did not associate with fibrinogen, d-dimer, platelet, hemoglobin or ANC.

In order to understand the relationship between IFNγ and clinical outcomes in MIS-C, we examined surrogate markers of severity. We used requirement of inotropes as a surrogate marker of severity. Surprisingly, patients within the IFNγ-high cluster had a lower likelihood of requiring inotropes than those in the IFNγ-low cluster (Figure 6F), implying that higher levels of IFNγ are associated with less severe illness in MIS-C patients. NTproBNP, a marker associated with cardiac damage, showed a similar trend, although the result was not statistically significant, likely due to a lack of power (Figure 6G). We performed this analysis on all patients with MIS-C and found that higher levels of NTproBNP were higher in patients requiring inotropes (Figure 6H). Thus, it appears higher levels of IFNγ may be associated with less severe illness in patients with MIS-C.

Previously, Vella et al. demonstrated that inotrope need was associated with higher frequencies of activation of CX3CR1+ CD8+ T-cells.^20^ CX3CR1 is expressed on CD4+ T-cells, monocytes, and effector-like CD8+ T-cells, and allows cells to interact with CX3CL1-expressing vasculature.^38, 39^ In MIS-C patients, activation of CX3CR1+ CD8+ T-cells (but not CX3CR1+ CD4+ T-cells; Supplemental Figure S4C) and IFNγ inversely correlated (Figure 6I; N=7, R=-0.83, p=0.02). However, levels of the CX3CR1 ligand, CX3CL1 had no relationship to CX3CR1+ CD8+ T-cell activation (Figure 6I). In our unbiased network analysis (Figure 6D) we identified CX3CL1 as an enriched protein despite it not reaching the thresholds used for our differential expression analysis (Figure 6C). When explicitly tested, we surpassingly observed an association between elevated CX3CL1 and IFNγ-high patients, a group of patients we demonstrated to be at lower risk for inotrope use (Figure 6J). The data suggest that free CX3CL1 levels in the plasma are independent of the frequency of CX3CR1+ CD8+ T-cell activation (Figure 6I), explaining their differential association with inotrope requirement. Thus, we demonstrate that patients with high IFNγ levels are less likely to require inotrope use and accordingly have lower levels of CX3CR1+ CD8+ T-cell activation.

## DISCUSSION

We used proteomic analysis to begin to unravel the complex and heterogeneous pathophysiology associated with MIS-C. We identify PLA2G2A as a candidate biomarker for MIS-C and show that PLA2G2A is associated with clinical features of TMA. We demonstrate that MIS-C patients are characterized by a disproportionately high CXCL9 response to IFNγ, implying a dysregulated response to IFNγ. We found that a subset of patients with MIS-C met modified criteria for the IFNγ-associated syndrome MAS. Patients with MIS-C showed heterogeneity based on IFNγ expression, and surprisingly, patients with higher IFNγ levels were less likely to require inotropes.

PLA2G2A is involved in host inflammatory responses and is associated with release of damage associated molecular pattern molecules (DAMPs) that play a key role in activation of the innate immune response.^40, 41^ PLA2G2A released from activated platelets is associated with neutrophilia. ^40^ Expression of PLA2G2A can be driven by IFNγ seceretion.^42–44^ In HSCT associated TMA, IFNγ has been identified as a key causative agent and part of an “interferon-complement loop” that contributes to vessel injury and TA-TMA pathophysiology.^45^

Overlapping phenotypes of MAS and TMA have been reported by others in other disease settings.^46^ We hypothesize that patients with MIS-C may be responding to IFNγ with excessive PLA2G2A production. However, this relationship is likely complex as patients with high IFNγ expression appear to be protected from cardiac toxicity associated with MIS-C. Whether or not there is a causative relationship between PLA2G2A and TMA or MIS-C will need to be determined in future experiments. Understanding the role of PLA2G2A in MIS-C pathophysiology is crucial because PLA2G2A and its associated pathways can be targeted with the common, inexpensive medication indomethacin.^47^ Furthermore, higher levels of PLA2G2A associate with clinical findings of TMA. Regardless of the cause of PLA2G2A expression, the levels are markedly high in almost all patients with MIS-C and differentiate MIS-C patients form other SARS-CoV-2 infected patients, identifying PLA2G2A as a candidate biomarker for MIS-C. Future studies will need to prospectively examine PLA2G2A in patients with MIS-C and acute SARS-CoV-2 infection and validate its role as a diagnostic biomarker of MIS-C.

Patients with MIS-C produced excessive CXCL9 in response to IFNγ. CXCL9 production has been associated with an increased risk of mortality in adult patients with severe COVID-19 disease and may contribute to cytokine storm in that setting.^48^ One potential explanation for the heightened CXCL9 response to IFNγ in MIS-C patients is lower levels of TRIM21, a protein notable for its ability to repress IFNγ signaling through degradation of interferon response factors.^34, 35^ TRIM21 is induced by IFNγ, and indeed was one of the upregulated factors in the IFNγ-high MIS-C group, suggesting that this negative feedback loop is at least still partially intact in MIS-C despite the generally lower levels of TRIM21.^49^ In keeping with IFNγ dysregulation, found that a subset of patients with MIS-C met modified criteria for MAS. We note that an IFNγ-high and -low subgrouping of MIS-C has been noted by others.^7^ While the connection to MAS was also invoked by Esteve-Solie et al., we have clearly shown a connection between IFNγ-high MIS-C patients and objective MAS features in our data.

Other features of MAS in patients with MIS-C have been reported in the literature, including overlapping cytokine phenotypes and hemophagocytosis on bone marrow aspirates.^50^ Although the evidence suggests that MIS-C does share many things in common with MAS, the clinical presentation of MIS-C patients is not fully consistent with a complete MAS phenotype and may be better understood as an occult MAS.^51, 52^ MAS is often treated with IL-1β blockade.^53^ Interestingly, IL-1β blockade with anakinra, a recombinant IL-1 receptor antagonist has been reported in many case reports as efficacious in MIS-C, and it is recommended for refractory patients in the American College of Rheumatology guidelines.^54^ We further note that IL-1RN, the endogenous IL-1 receptor antagonist is upregulated in the IFNγ high group (Figure 6B). It is interesting to speculate whether this confers some of the cardiac protection seen in this group.

Curiously, we found that IFNγ-high patients were less likely to require inotropes, a result that is not consistent with other disorders associated with dysregulated IFNγ production.^28^ This result was confirmed by an anti-correlation between IFNγ and CD8+ CX3CR1+ T-cells, a cell population which has been previously associated with an increased need for inotropes in MIS-C patients.^20^ The novel observation that IFNγ may predict cardiac pathology in MIS-C may prove to be useful clinically as cytokine testing becomes more readily available in hospital settings. Future work should test this prospectively.

We have made several key observations about the underlying pathophysiology of MIS-C, however, our study is limited by several factors. First, we retrospectively applied clinical categorizations for TMA and MAS to patients included in this study in order to understand relationships between clinical phenotypes and cytokine signatures. Neither of the clinical criteria used have been validated in this setting, and future studies should prospectively validate clinical criteria for these disorders in pediatric patients. Due to the rarity of this condition, we are underpowered to examine the full scope of clinical heterogeneity among MIS-C patients. We have identified important pathways to be examined in further detail in future mechanistic studies.

MIS-C is an important emergent syndrome characterized by excessive cytokine production and vascular-endothelial dysfunction. We have demonstrated that PLA2G2A is a key marker of MIS-C and associates with TMA. We have uncovered dysregulations in IFNγ responses in MIS-C patients and that IFNγ levels themselves reveal clinically relevant heterogeneity within MIS-C. Future studies should mechanistically examine the relationships between IFNγ, PLA2G2A, and their drivers in MIS-C initiation.

## ONLINE METHODS

### Study Approvals

This study was conducted in accordance with the Declaration of Helsinki and received approval from the Institutional Review Board (IRB) at CHOP. Verbal consent for this minimal risk study was obtained from patients or their legally authorized representative. Consent forms were signed by the consenting study team member and a copy was provided to the study participant or legally authorized representative. If appropriate, assent was obtained from children who were 7 years of age or older.

For remnant samples obtained from healthy controls, protected health information (PHI) was not recorded. A limited chart review of this cohort was granted by the CHOP IRB to determine that patients met criteria to be considered “healthy”. CHOP IRB granted exemption criteria per 45 CFR 46.104(d) 4(ii) and waiver of HIPPA authorization.

### Study Design and Population

Patients were eligible to be prospectively enrolled in the Children’s Hospital of Philadelphia (CHOP) SARS-CoV-2 biobank if they were admitted to CHOP during the COVID-19 pandemic and had either a positive SARS-CoV-2 reverse transcriptase polymerase chain reaction (RT-PCR) result from upper respiratory tract mucosa, had a positive SARS-CoV-2 antibody or met criteria for MIS-C. These enrollment criteria have been described in detail previously.^17, 21^

### Sample Collection

Patient samples were collected as soon as possible after admission to the hospital. All samples were collected in combination with a clinical blood draw. Blood for protein analysis was collected in a lithium heparin tube and separated into plasma and cell pellets. Components were frozen and stored in liquid nitrogen and -80 freezer respectively. Batch analysis was performed on plasma samples. Blood for flow cytometry was collected in a sodium heparin tube.^20^ In patients who remained in hospital, samples were collected weekly.

Plasma from otherwise healthy control patients who were being evaluated for symptoms of a bleeding disorder (such as epistaxis) were obtained from discarded plasma from the coagulation lab at CHOP. A limited chart review was performed to confirm these patients had no comorbid medical illnesses. Patients found to have comorbid medical issues other than a possible isolated bleeding disorder were excluded.

### Data Collection

Clinical and laboratory data were abstracted from electronic patient charts on to standardized case report forms created using the Research Electronic Data Capture database (REDCap; Vanderbilt University, Nashville TN USA).^55^ Data were abstracted by a clinician or clinical research assistant. All data elements were validated by a physician.

### Clinical categorization

Enrolled patients were prospectively classified in to 3 groups by physicians with expertise pediatric in hematology/oncology (CD, DT), pediatric infectious diseases (HB) or pediatric rheumatology (EB). Patient classification criteria have been described in detail previously.^17^ In brief, Severe COVID-19 was defined as patients requiring new mechanical or non-invasive ventilatory support, or an increase in respiratory support above their baseline requirement. Minimal COVID-19 was defined as either an incidental finding of SARS-CoV-2 positivity during testing prior to an admission, diagnostic test or procedure or mild symptoms due to COVID-19 that did not require non-invasive mechanical ventilation. Patients who required low flow oxygen alone were categorized as Minimal COVID-19. MIS-C was defined per the Centers for Disease Control (CDC) criteria as fever, evidence of inflammation (elevated CRP, ESR or procalcitonin), multisystem organ involvement with at least two organ systems (cardiac, renal, respiratory, hematologic, dermatologic, gastrointestinal or neurologic), evidence of past or current positive SARS-CoV-2 infection by RT-PCR, serology or proven exposure to a close contact within four weeks prior to the onset of symptoms.^56^

Patients were categorized as having thrombotic microangiopathy (TMA) based on criteria reported previously.^17^ Only patients on whom a Hematoxylin-Eosin stained peripheral blood smear available were evaluated for TMA (N=34). The criteria used included presence of thrombocytopenia, microangiopathic hemolytic anemia and organ dysfunction. Hemolytic anemia was defined as anemia for age and schistocytes present on a peripheral blood smear.

Organ dysfunction was defined as cardiac dysfunction (troponin greater than the upper limit of normal for age or requirement of inotropes), renal dysfunction (based on the Kidney Disease: Improving Global Outcomes (KDIGO) criteria)^57^ or liver dysfunction (bilirubin greater than twice the upper limit of normal for age, alanine aminotransferase or aspartate aminotransferase greater than three times the upper limit of normal).

In patients on whom a ferritin was available (N=40), a modified version of the Ravelli criteria for MAS were applied.^32^ We modified the Ravelli criteria to exclude triglyceride levels because they were not measured in the majority of our patients. We defined a modified Ravelli criteria as ferritin>684ng/mL and any 2 of 1) platelet count<181,000/uL, AST>48U/L, or fibrinogen<360mg/dL. If any data were missing, they were imputed as negative/normal in order to bias our categorization towards the null hypothesis.

### Proteomic analysis

Plasma protein levels were analyzed using the Olink Explore 1536/384 panel (Olink Proteomics, Uppsala, Sweden). Data were reported in normalized protein expression values (NPX). NPX is an arbitrary unit in a Log2 scale calculated from inverted, normalized Ct values. All assay validation data are available on the Olink website (www.olink.com).

### sC5b9 ELISA Assay

sC5b9 levels were determined using enzyme-linked immunosorbent assays (ELISA; Cat. #558315; BD Biosciences, San Jose CA, USA). Assays were performed at two dilutions; all samples were assayed in triplicate, using manufacturer protocols. As previously described, we set the upper limit of normal for our sC5b9 assay at 247 ng/mL. sC5b9 measurements were row normalized to a Z-score and log2 transformed in order to be comparable to NPX scores.

### Flow cytometry analysis

Detailed methods for flow cytometry analysis have been described previously.^20^ Briefly, peripheral blood mononuclear cells (PBMCs) were isolated from blood collected in sodium heparin tubes. PBMCs were stained with a live/dead stain and then treated with Fc block before staining for chemokine receptors, surface markers and intracellular markers. Samples were acquired on a 5 laser FACS Symphony A5 (BD Biosciences). All analysis was performed in FlowJo (Treestar, version 10.6.2). Flow cytometry data were transformed to a Z-score for all of the values for each gate used in order to be able to compare to the Log2 transformed NPX scores used in the Olink dataset.

### General statistical methods

All statistical analyses were performed in R (version 4.0.4) using RStudio (RStudio, PBC, Boston, MA).^58^ Data were assumed to be non-parametric unless normality was demonstrated. Correlations were performed with Spearman’s rank correlation coefficient for non-parametric data and Pearson’s correlation coefficient for parametric data. Kruskall-Wallis testing was performed to compare three or more groups and Wilcoxcon signed-rank test was performed for paired groups. To test for associations between discrete variables, Fisher’s exact test was used if small numbers were available in each cell. Otherwise, Chi-square testing was used. Unless otherwise stated, significance was based on a fold change greater than 2 or less the -2 and a false discovery rate (FDR; using Benjamini-Hochberg correction) value of less than 0.01.

### Clustering analysis

The T-distributed Stochastic Neighbor Embedding (tSNE) for R package was used to perform tSNE clustering.^59^ Principal component analysis was performed using R version 4.0.4. The factoextra package (https://cran.r-project.org/web/packages/factoextra/index.html) was used to extract and visualize PCA elements.

### Pathway analysis

We completed a differential expression analysis of all proteins using a log2fold change threshold of 2 and FDR threshold of 0.01. Proteins from the Olink data set were inputted in to PathfindR along with their respective log2fold change and FDR value.^25^ Using an active subnetwork enrichment analysis approach, PathfindR outputs a table that represents enriched pathways identified from the protein list inputted into the program. We used the KEGG pathway database (https://www.genome.jp/kegg/pathway.html) which includes a manual collection of pathway maps examining a total of 777,729 molecular pathways, with 544 main pathways included. Only pathways that had a p-value of <=0.01 were considered. The final table produced by PathfindR includes a table of significant pathways with an associated adjusted p-value, a fold enrichment value of the pathway, the lowest and highest p-values generated from each iteration of the pathways analysis, and the upregulated and downregulated proteins from the input protein list for every pathway.^25^

## Data Availability

The authors confirm that the data supporting the findings of this study are available within the article and its supplementary materials.

## ACKNOWLEDGEMENTS

We gratefully acknowledge the support of the Children’s Hospital of Philadelphia Frontiers program and the assistance of Mr. Jansen Weaver, Mr. Daniel Fields and Mr. Kienan O’Brien.

## AUTHOR CONTRIBUTIONS

EMB, DTT, and HB equally contributed to this work and are co-senior authors. CD, SEH, LV, KES, MPL, EJW, EB, HB, and DTT designed the study. RS, FB, LV, JRG, AEB, DO, AF, CD, SC, EMB analyzed data. CB, JRG, AEB performed experiments. CD, JL, TL and KOM enrolled subjects. CD, SH, KOM, JL, LV, TL, DTT, HB and EMB abstracted clinical data. All authors contributed intellectually and reviewed and revised the manuscript.

## References

1. Verdoni L, et al. An outbreak of severe Kawasaki-like disease at the Italian epicentre of the SARS-CoV-2 epidemic: an observational cohort study. The Lancet. 2020;(Published online May 13, 2020).

2. Riphagen S, et al. Hyperinflammatory shock in children during COVID-19 pandemic. The Lancet. 2020;(published online May 7, 2020).

3. Viner RM, Whittaker E. Kawasaki-like disease: emerging complication during the COVID-19 pandemic. The Lancet. 2020;(Published online May 13, 2020).

4. Feldstein LR, et al. Multisystem Inflammatory Syndrome in U.S. Children and Adolescents. N Engl J Med. 2020;383(4):334–346.

5. Consiglio CR, et al. The Immunology of Multisystem Inflammatory Syndrome in Children with COVID-19. Cell. 2020;183(4):968–981.e967.

6. Rowley AH. Understanding SARS-CoV-2-related multisystem inflammatory syndrome in children. Nat Rev Immunol. 2020;20(8):453–454.

7. Esteve-Sole A, et al. Similarities and differences between the immunopathogenesis of COVID-19-related pediatric multisystem inflammatory syndrome and Kawasaki disease. J Clin Invest. 2021;131(6).

8. Ouldali N, et al. Association of Intravenous Immunoglobulins Plus Methylprednisolone vs Immunoglobulins Alone With Course of Fever in Multisystem Inflammatory Syndrome in Children. JAMA. 2021;325(9):855–864.

9. Elias MD, et al. Management of Multisystem Inflammatory Syndrome in Children Associated With COVID-19: A Survey From the International Kawasaki Disease Registry. CJC Open. 2020;2(6):632–640.

10. McMurray JC, et al. Multisystem Inflammatory Syndrome in Children (MIS-C), a Post-viral Myocarditis and Systemic Vasculitis-A Critical Review of Its Pathogenesis and Treatment. Front Pediatr. 2020;8:626182.

11. Brodsky NN, et al. The Mystery of MIS-C Post-SARS-CoV-2 Infection. Trends Microbiol. 2020;28(12):956–958.

12. Gruber CN, et al. Mapping Systemic Inflammation and Antibody Responses in Multisystem Inflammatory Syndrome in Children (MIS-C). Cell. 2020;183(4):982–995.e914.

13. Lee PY, et al. Distinct clinical and immunological features of SARS-CoV-2-induced multisystem inflammatory syndrome in children. J Clin Invest. 2020(1558–8238 (Electronic)).

14. Fajgenbaum DC, June CH. Cytokine Storm. N Engl J Med. 2020;383(23):2255–2273.

15. Carter MJ, et al. Peripheral immunophenotypes in children with multisystem inflammatory syndrome associated with SARS-CoV-2 infection. Nat Med. 2020;26(11):1701–1707.

16. Klok FA, et al. Confirmation of the high cumulative incidence of thrombotic complications in critically ill ICU patients with COVID-19: An updated analysis. Thromb Res. 2020;191:148–150.

17. Diorio C, et al. Evidence of thrombotic microangiopathy in children with SARS-CoV-2 across the spectrum of clinical presentations. Blood Adv. 2020;4(23):6051–6063.

18. George JN, Nester CM. Syndromes of Thrombotic Microangiopathy. N Engl J Med. 2014;371(7):654–666.

19. Dvorak CC, et al. Transplant-Associated Thrombotic Microangiopathy in Pediatric Hematopoietic Cell Transplant Recipients: A Practical Approach to Diagnosis and Management. Front Pediatr. 2019;7:133.

20. Vella L, et al. Deep Immune Profiling of MIS-C demonstrates marked but transient immune activation compared to adult and pediatric COVID-19. Sci Immunol. 2020;6(57).

21. Diorio C, et al. Multisystem inflammatory syndrome in children and COVID-19 are distinct presentations of SARS-CoV-2. J Clin Invest. 2020;130(11):5967–5975.

22. Anderson EM, et al. SARS-CoV-2 antibody responses in children with MIS-C and mild and severe COVID-19. J Pediatric Infect Dis Soc. 2020.

23. Diorio C, et al. Convalescent plasma for pediatric patients with SARS-CoV-2-associated acute respiratory distress syndrome. Pediatr Blood Cancer. 2020;67(11):e28693.

24. Hall C. Essential biochemistry and physiology of (NT-pro)BNP. Eur J Heart Fail. 2004;6(3):257–260.

25. Ulgen E, et al. pathfindR: An R Package for Comprehensive Identification of Enriched Pathways in Omics Data Through Active Subnetworks. Front Genet. 2019;10:858.

26. Aste-Amezaga M, et al. Molecular mechanisms of the induction of IL-12 and its inhibition by IL-10. J Immunol. 1998;160(12):5936–5944.

27. Mizuta M, et al. Clinical significance of serum CXCL9 levels as a biomarker for systemic juvenile idiopathic arthritis associated macrophage activation syndrome. Cytokine. 2019;119:182–187.

28. Crayne CB, et al. The Immunology of Macrophage Activation Syndrome. Front Immunol. 2019;10(119).

29. Bracaglia C, et al. Elevated circulating levels of interferon-γ and interferon-γ-induced chemokines characterise patients with macrophage activation syndrome complicating systemic juvenile idiopathic arthritis. Ann Rheum Dis. 2017;76(1):166–172.

30. Grom AA, et al. Macrophage activation syndrome in the era of biologic therapy. Nat Rev Rheumatol. 2016;12(5):259–268.

31. Yuan S, et al. Serum soluble VSIG4 as a surrogate marker for the diagnosis of lymphoma-associated hemophagocytic lymphohistiocytosis. Br J Haematol. 2020;189(1):72–83.

32. Ravelli A, et al. 2016 Classification Criteria for Macrophage Activation Syndrome Complicating Systemic Juvenile Idiopathic Arthritis. Ann Rheum Dis. 2016;75:481–489.

33. Schulert GS, et al. Monocyte and bone marrow macrophage transcriptional phenotypes in systemic juvenile idiopathic arthritis reveal TRIM8 as a mediator of IFN-γ hyper-responsiveness and risk for macrophage activation syndrome. Ann Rheum Dis. 2020.

34. Higgs R, et al. Self protection from anti-viral responses--Ro52 promotes degradation of the transcription factor IRF7 downstream of the viral Toll-Like receptors. PLoS One. 2010;5(7):e11776.

35. Yoshimi R, et al. Gene disruption study reveals a nonredundant role for TRIM21/Ro52 in NF-kappaB-dependent cytokine expression in fibroblasts. J Immunol. 2009;182(12):7527–7538.

36. Ma L, et al. Increased complement activation is a distinctive feature of severe SARS-CoV-2 infection. bioRxiv. 2021.

37. McGeachy MJ, et al. The IL-17 Family of Cytokines in Health and Disease. Immunity. 2019;50(4):892–906.

38. Imai T, et al. Identification and molecular characterization of fractalkine receptor CX3CR1, which mediates both leukocyte migration and adhesion. Cell. 1997;91(4):521–530.

39. Umehara H, et al. Fractalkine in Vascular Biology. Arterioscler Thromb Vasc Biol. 2004;24(1):34–40.

40. Boudreau LH, et al. Platelets release mitochondria serving as substrate for bactericidal group IIA-secreted phospholipase A2 to promote inflammation. Blood. 2014;124(14):2173–2183.

41. Kitano N, et al. Patient Age and the Seasonal Pattern of Onset of Kawasaki’s Disease. N Engl J Med. 2018;378(21):2048–2049.

42. Lindbom J, et al. Interferon gamma-induced gene expression of the novel secretory phospholipase A2 type IID in human monocyte-derived macrophages is inhibited by lipopolysaccharide. Inflammation. 2005;29(2-3):108–117.

43. Ponzoni M, et al. Stimulation of receptor-coupled phospholipase A2 by interferon-gamma. FEBS Lett. 1992;310(1):17–21.

44. Wu T, et al. Interferon-gamma induces the synthesis and activation of cytosolic phospholipase A2. J Clin Invest. 1994;93(2):571–577.

45. Jodele S, et al. Interferon-complement loop in transplant-associated thrombotic microangiopathy. Blood Adv. 2020;4(6):1166–1177.

46. Gloude NJ, et al. Thinking Beyond HLH: Clinical Features of Patients with Concurrent Presentation of Hemophagocytic Lymphohistiocytosis and Thrombotic Microangiopathy. J Clin Immunol. 2020;40(5):699–707.

47. Berry E, et al. Matrix metalloproteinase-2 negatively regulates cardiac secreted phospholipase A2 to modulate inflammation and fever. J Am Heart Assoc. 2015;4(4).

48. Abers MS, et al. An immune-based biomarker signature is associated with mortality in COVID-19 patients. JCI Insight. 2021;6(1):11.

49. Sjöstrand M, et al. Expression of the immune regulator tripartite-motif 21 is controlled by IFN regulatory factors. J Immunol. 2013;191(7):3753–3763.

50. Lacinel Gurlevik S, et al. Hemophagocytosis in bone marrow aspirates in multisystem inflammatory syndrome in children. Pediatr Blood Cancer. 2021:e28931.

51. Behrens EM, et al. Occult macrophage activation syndrome in patients with systemic juvenile idiopathic arthritis. J Rheumatol. 2007;34(5):1133–1138.

52. Bleesing J, et al. The diagnostic significance of soluble CD163 and soluble interleukin-2 receptor alpha-chain in macrophage activation syndrome and untreated new-onset systemic juvenile idiopathic arthritis. Arthritis Rheumatol. 2007;56(3):965–971.

53. Mehta P, et al. Silencing the cytokine storm: the use of intravenous anakinra in haemophagocytic lymphohistiocytosis or macrophage activation syndrome. Lancet Rheumatol. 2020;2(6):e358–e367.

54. Henderson LA, et al. American College of Rheumatology Clinical Guidance for Multisystem Inflammatory Syndrome in Children Associated With SARS-CoV-2 and Hyperinflammation in Pediatric COVID-19: Version 2. Arthritis Rheumatol. 2021;73(4):e13–e29.

55. Harris PA, et al. The REDCap consortium: Building an international community of software platform partners. J Biomed Inf. 2019;95:103208.

56. CDC. Multisystem Inflammatory Syndrome in Children (MIS-C) Associated with Coronavirus Disease 2019 (COVID-19); 2020.

57. Kaddourah A, et al. Epidemiology of Acute Kidney Injury in Critically Ill Children and Young Adults. N Engl J Med. 2016;376(1):11–20.

58. R Studio Team. RStudio: Integrated Development for R. Boston, MA: PBC; 2020.

59. Van der Maaten L, Hinton G. Visualizing data using t-SNE. J Mach Learn Res. 2008;9(11):2579–2605.

