## Supplementary figures and images for "Proteomic Profiling of MIS-C Patients Reveals Heterogeneity Relating to Interferon Gamma Dysregulation and Vascular Endothelial Dysfunction"

### Supplemental Figure 1

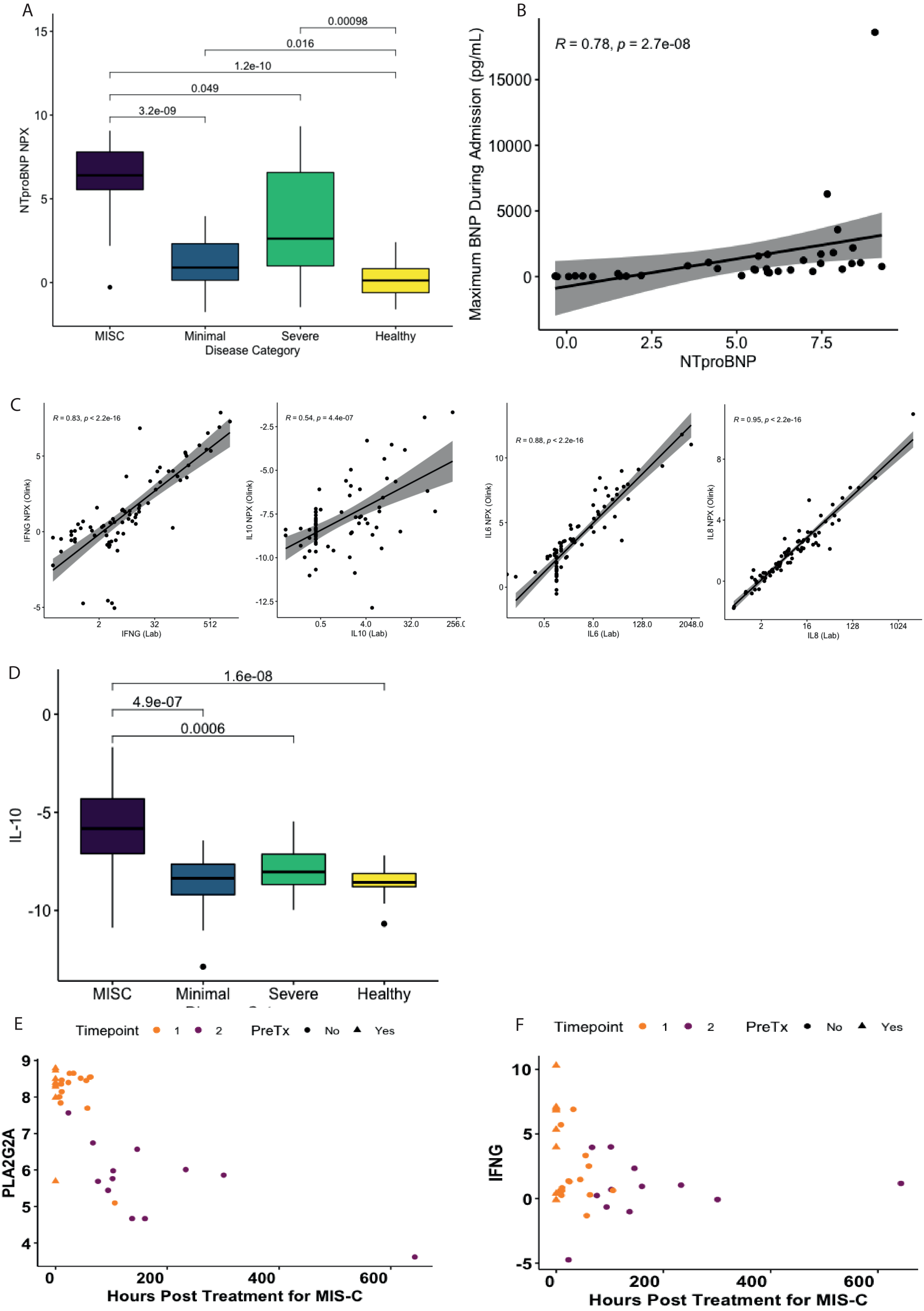

### Supplemental Figure 2

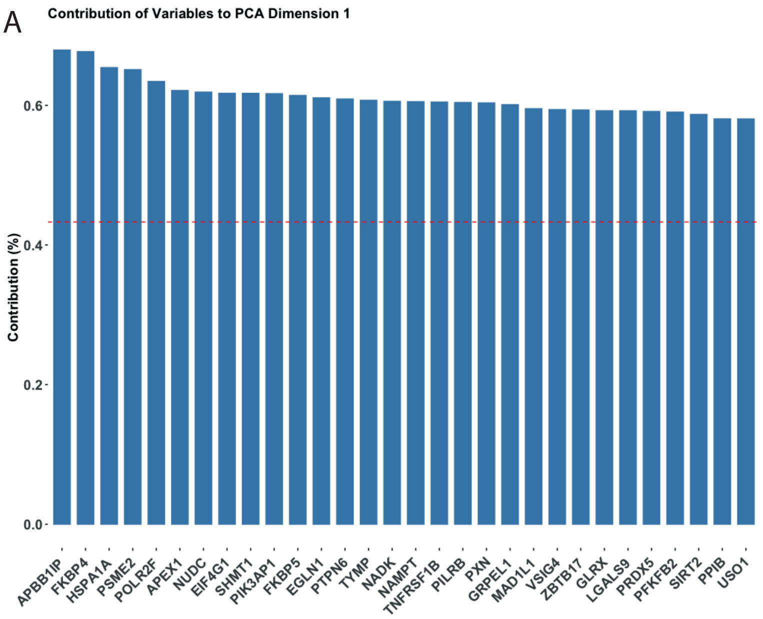

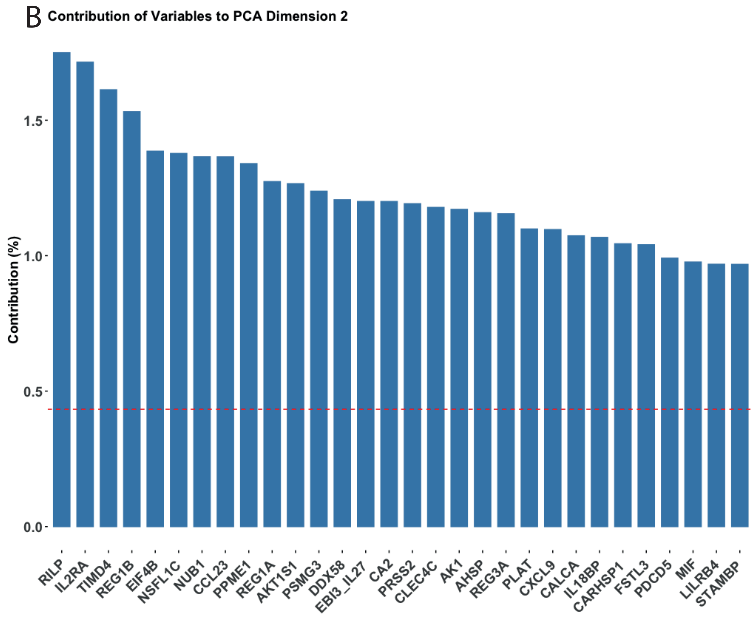

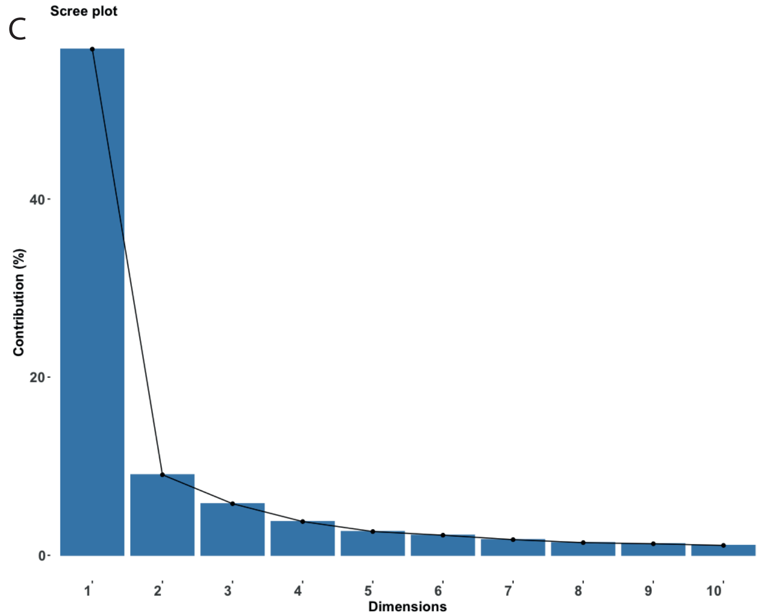

### Supplemental Figure 3

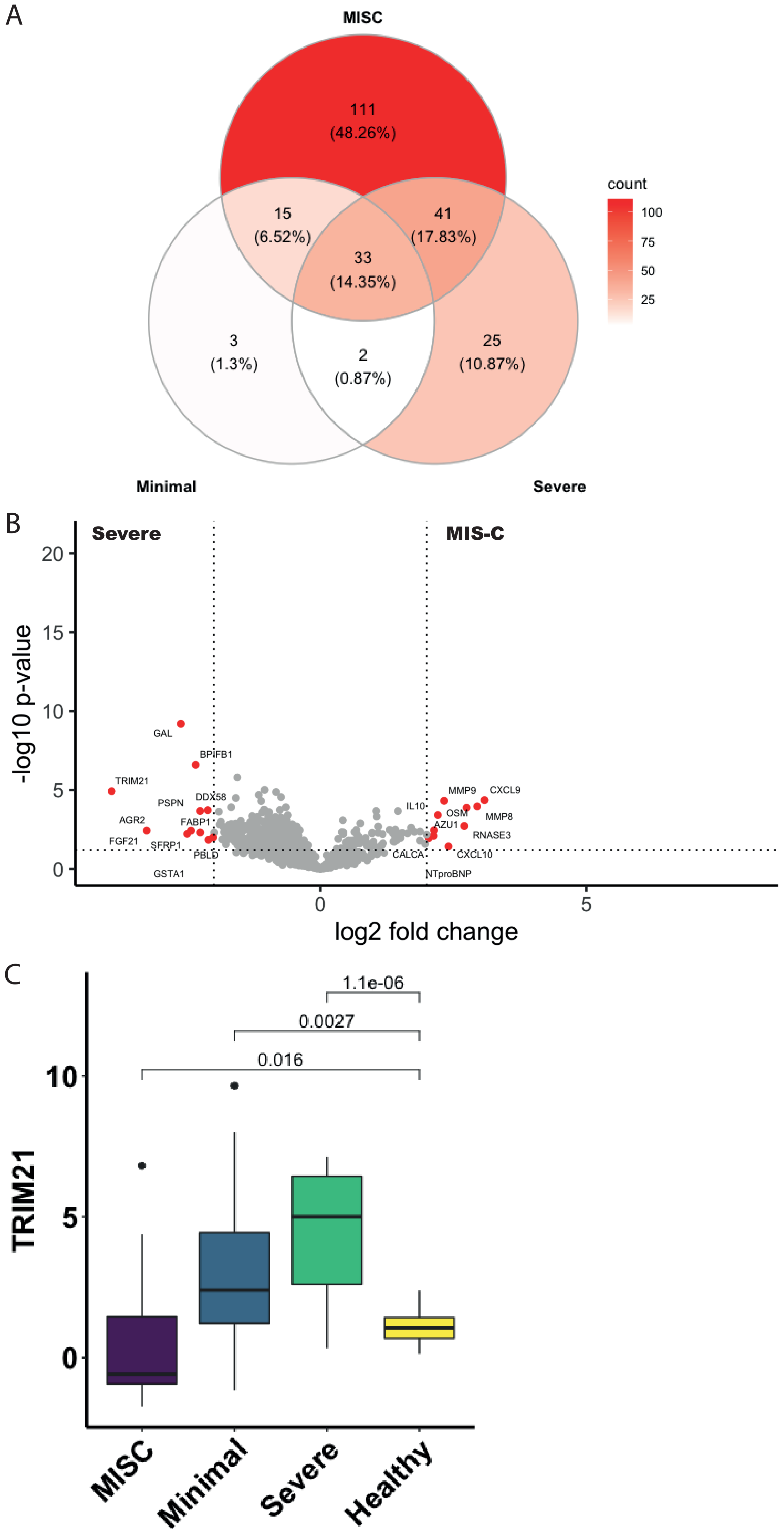
