## Supplemental Tables for "Proteomic Profiling of MIS-C Patients Reveals Heterogeneity Relating to Interferon Gamma Dysregulation and Vascular Endothelial Dysfunction"

Supplemental Table S1. Demographic and clinical information about patients with MIS-C, severe COVID-19 or minimal COVID-19.

Supplemental Table S2. Most extreme laboratory values during admission for patients with MIS-C, severe COVID-19 or minimal COVID-19.

Supplemental Table S3. Contingency Table of MAS by Disease Category.

Supplemental Table S4. Contingency Table of TMA by Disease Category.

Supplemental Table S5. Contingency tables for MIS-C patients in the IFNγ-high versus -low clusters and those who met criteria for TMA or MAS.

Supplemental Table S1. Demographic and clinical information about patients with MIS-C, severe COVID-19 or minimal COVID-19.

|  | MISC  N =22 | Severe  COVID-19  N =15 | Minimal COVID-19  N = 26 |
| --- | --- | --- | --- |
| Age Median (IQR) | 9 (6.25-13) | 16 (14.5-18) | 14 (5.75-17) |
| BMI Percentile Median (IQR) | 92.2 (80-97.5; N=21) | 81.6 (66.1, 94.8; N=14) | 73.4 (47.2, 93.8; N= 22) |
| Gender N (%) |  |  |  |
| *Female* | 10 (45) | 7 (47) | 12 (46) |
| *Male* | 12 (55) | 8 (53) | 14 (54) |
| Race N (%) |  |  |  |
| *White* | 8 (36) | 5 (33) | 11 (42) |
| *Black* | 11 (50) | 5 (33) | 12 (46) |
| *Other* | 2 (9) | 4 (27) | 3 (12) |
| *Declined* | 1 (5) | 1 (7) | 0 (0) |
| Ethnicity N (%) |  |  |  |
| *Hispanic (1)* | 3 (14) | 4 (27) | 4 (15) |
| *Not Hispanic (2)* | 18 (82) | 10 (67) | 22 (85) |
| *Declined (99)* | 1 (4) | 1 (6) | 0 (0) |
| PICU Admission N (%) |  |  |  |
| *Yes* | 17 (77) | 15 (100) | 21 (81) |
| *No* | 5 (23) | 0 | 5 (19) |
| ECMO N (%) |  |  |  |
| *Yes* | 0 | 2 (13) | 0 |
| *No* | 22 (100) | 13 (87) | 26 (100) |
| Inotropic Support N (%) |  |  |  |
| *Yes* | 7 (32) | 7 (47) | 0 |
| *No* | 15 (68) | 8 (53) | 26 (100) |
| Lymphopenic During Admission N (%) |  |  |  |
| *Yes* | 18 (82) | 9 (60) | 14 (54) |
| *No* | 4 (18) | 6 (40) | 12 (46) |
| Neutropenic During Admission N (%) |  |  |  |
| *Yes* | 0 | 5 (33) | 8 (31) |
| *No* | 22 (100) | 10 (67) | 18 (69) |
| Previously Healthy N (%) |  |  |  |
| *Yes* | 22 (100) | 2 (13) | 5 (19) |
| *No* | 0 | 13 (87) | 21 (81) |

Supplemental Table S2. Most extreme laboratory values during admission for patients with MIS-C, severe COVID-19 or minimal COVID-19.

|  | MIS-C | | Severe COVID-19 | | | Minimal COVID-19 | |
| --- | --- | --- | --- | --- | --- | --- | --- |
| Value (reference range) | Median (IQR) | N | Median (IQR) | | N | Median (IQR) | N |
| COAGULATION | | | | | | | |
| D-Dimer, highest  (0.27 - 0.60 μg/ml FEU) | 5.01 (4.01, 6.52) | 21 | 1.06 (0.63, 5.4) | | 14 | 0.98 (0.583, 1.660 | 6 |
| PT, highest (11.6-13.8 secs) | 14.1 (14.5, 17.3) | 22 | 14.7 (13, 17.9) | | 14 | 13.3 (12.3, 14.3) | 13 |
| PTT, highest (22-36 secs) | 31.8 (29.5, 35.2) | 21 | 47.2 (35.8, 57.9) | | 14 | 29.7 (27.1, 33.7) | 13 |
| Fibrinogen, lowest (172-471 mg/dL) | 297 (230, 394) | 21 | 303 (269, 478) | | 12 | 407 (262, 734) | 6 |
| CHEMISTRY | | | | | | | |
| LDH, highest (360-730 U/L) | 767 (600,920) | 21 | 966 (785,3160) | | 8 | 717 (575, 891) | 6 |
| AST, highest (15-45 U/L) | 82.0 (66.0, 104) | 22 | 115 (69.5, 293) | | 15 | 59.0 (40.0, 72.0) | 21 |
| ALT, highest (10-35 U/L) | 62.0 (38.0, 87.8) | 22 | 54.0 (32.5, 134) | | 15 | 30.0 (23.0, 47.0) | 21 |
| Creatinine, highest  (0.3-0.8 mg/dL) | 0.600 (0.525, 1.20) | 22 | 0.5 (0.300, 1.20) | | 15 | 0.450 (0.300, 0.725) | 24 |
| Bilirubin, highest (0.6-1.4 mg/dL) | 0.950 (0.600, 1.3) | 22 | 0.700 (0.50, 0.95) | | 15 | 0.700 (0.400, 1.00) | 21 |
| Sodium, lowest (136-145 mmol/L) | 131 (130, 135) | 22 | 136 (133, 138) | | 15 | 136 (133, 137) | 24 |
| HEMATOLOGY | | | | | | | |
| Neutrophils (1,540 - 7,040 /uL) |  |  |  | |  |  |  |
| *Lowest* | 7330 (5110, 8150) | 22 | 2420 (1140, 3770) | | 11 | 3090 (1490, 3900) | 26 |
| *Highest* | 13500 (11200, 21400) | 22 | 7970 (6200, 13000) | | 11 | 5250 (3030, 8930) | 26 |
| Lymphocytes, lowest, (970-3,260 /uL) | 610 (305, 878) | 22 | 890 (390, 1380) | | 11 | 1220 (585, 2100) | 26 |
| Hemoglobin, lowest  (12-16 g/dL) | 8.45 (7.53, 9.53) | 22 | 10.1 (7.30, 11.9) | | 11 | 9.75 (7.83, 12.1) | 26 |
| Platelets, lowest (150-400 K/μL) | 150 (124, 189) | 22 | 146 (86.5, 175) | | 15 | 208 (128, 291) | 26 |
| INFLAMMATORY & CARDIAC | | | | | | | |
| Ferritin, highest (10.0-82.0 ng/ml) | 892 (665, 1370 | 21 | 217 (165, 1140) | | 13 | 1060 (307, 1470) | 6 |
| CRP, highest (0-0.9 mg/dL) | 28.2 (19.2, 35.1 | 22 | 16.1 (5.75, 31.7 | | 15 | 4.55 (1.30, 18.2) | 16 |
| ESR, highest (0-20 mm/hr) | 68.0 (46.0, 103) | 21 | 20.0 (13.0, 23.0) | 5 | | 46.5 (13.8, 85.5) | 12 |
| BNP, highest (≤100 pg/mL) | 1060 (563, 1720) | 21 | 223 (50.2, 454) | | 11 | 25.8 (10.0, 239) | 4 |
| Troponin, highest  (<0.3 ng/ml) | 0.600 (0.07, 1.87) | 21 | 0.115 (0.02, 0.93) | | 12 | 0.01 (0.01, 0.015) | 3 |

Supplemental Table S3 Contingency Table of MAS by Disease Category.

|  | MAS | No MAS |
| --- | --- | --- |
| MIS-C | 11 | 10 |
| Minimal | 3 | 3 |
| Severe | 9 | 4 |

Supplemental Table S4 Contingency Table of TMA by Disease Category.

|  | TMA | No TMA |
| --- | --- | --- |
| MISC | 7 | 8 |
| Severe | 5 | 3 |
| Minimal | 1 | 10 |

Supplemental Table S5. Contingency tables for MIS-C patients in the IFNγ-high versus -low clusters and those who met criteria for TMA or MAS.

TMA

|  | IFNγ-low | IFNγ-high |
| --- | --- | --- |
| No TMA | 5 | 3 |
| TMA | 6 | 1 |
| %TMA | 55% | 25% |

MAS

|  | IFNγ-low | IFNγ-high |
| --- | --- | --- |
| No MAS | 8 | 1 |
| MAS | 5 | 5 |
| %MAS | 38% | 83% |
