## Supplemental Figure Legends for "Proteomic Profiling of MIS-C Patients Reveals Heterogeneity Relating to Interferon Gamma Dysregulation and Vascular Endothelial Dysfunction"

Supplemental Figure List

**Supplemental Figure S1. Validations of the Olink data set compared to previously published datasets.** (A) N-terminal prohormone brain natriuretic peptide (NTproBNP) levels compared between patients with Multisystem Inflammatory Syndrome in Children (MIS-C; N=22), minimal SARS-CoV-2 infection (N=26), severe COVID-19 (N=15) and healthy controls (N=25). P-values computed with pairwise comparisons using Wilcoxon rank sum test following Kruskal-Wallis testing. Boxes and whiskers show median and interquartile range. (B) In patients on whom a brain-type natriuretic peptide (BNP) was measured during admission (N=36), these values were correlated with NTproBNP with a strong correlation. R values computed with Spearman correlation. (C) Correlations between interferon gamma (IFNγ), interleukin-10 (IL-10), IL-6 and IL-8 measured by the Olink data set and by the clinical lab. R values computed with Spearman correlation. (D) IL-10 levels between the four disease categories. P-values computed with pairwise comparisons using Wilcoxon rank sum test following Kruskal-Wallis testing. Decay in phospholipase A2 (PLA2G2A; E) and IFNγ; (F) over time for MIS-C patients is shown (N=22). Dots are colored by order of draw (first versus second timepoint). Shapes represent if samples were drawn prior to or after treatment with intravenous immune globulin (IVIG) or corticosteroids.

**Supplemental Figure S2. Components of Principal Component Analysis (PCA) in Figure 2B.** Top 30 proteins that contribute to PCA Dimension 1 (A) and PCA Dimension 2 (B). Scree plot of eigenvalues of principal components of the PCA is shown in panel (C).

**Supplemental Figure S3. Differentially expressed proteins between different disease states.** (A) Venn diagram of overlap of differentially expressed proteins between different disease states and healthy patients. (B) Differentially expressed proteins between patients with Multisystem Inflammatory Syndrome in Children (MIS-C; N=22) and severe COVID-19 (N=15). Red dots represent proteins with a nominal p-value of less than 0.05 and a log2fold change of greater than 2. (C) TRIM21 levels among patients with MIS-C (N=22), Severe (N=15), Minimal disease (N=26) and healthy controls (N=25). P-values computed with pairwise comparisons using Wilcoxon rank sum test following Kruskal-Wallis testing. Boxes and whiskers show median and interquartile range.

**Supplemental Figure S4. Heterogeneity of disease phenotypes among MIS-C patients.** In panels (A) t-distributed Stochastic Neighbor Embedding (tSNE) plot from Figure 1A are shown with dots colored by whether or not patients met criteria for thrombotic microangiopathy (TMA), macrophage activation syndrome (MAS), neither phenotype or both phenotypes. Rings around dots are colored by disease category, including Multisystem Inflammatory Syndrome in Children (MIS-C; N=22), minimal SARS-CoV-2 infection (N=26), severe COVID-19 (N=15) and healthy controls (N=25). Panel (B) shows a similar output but with internal dots colored by interferon gamma (IFNγ) expression. Panel (C) demonstrates a lack of correlation between IFNγ level and percent DR+CD38+ non-naïve CD4+ CX3CR1+ T-cells in MIS-C patients (N=7). Dots colored by CX3CL1 expression. R value computed using Pearson’s correlation coefficient after normality was demonstrated.
